# Supervised Image Classification Algorithm Using Representative Spatial Texture Features: Application to COVID-19 Diagnosis Using CT Images

**DOI:** 10.1101/2020.12.03.20243493

**Authors:** Zehor Belkhatir, Raúl San José Estépar, Allen R. Tannenbaum

## Abstract

Although there is no universal definition for texture, the concept in various forms is nevertheless widely used and a key element of visual perception to analyze images in different fields. The present work’s main idea relies on the assumption that there exist representative samples, which we refer to as references as well, i.e., “good or bad” samples that represent a given dataset investigated in a particular data analysis problem. These representative samples need to be accounted for when designing predictive models with the aim of improving their performance. In particular, based on a selected subset of texture gray-level co-occurrence matrices (GLCMs) from the training cohort, we propose new representative spatial texture features, which we incorporate into a supervised image classification pipeline. The pipeline relies on the support vector machine (SVM) algorithm along with Bayesian optimization and the Wasserstein metric from optimal mass transport (OMT) theory. The selection of the best, “good and bad,” GLCM references is considered for each classification label and performed during the training phase of the SVM classifier using a Bayesian optimizer. We assume that sample fitness is defined based on closeness (in the sense of the Wasserstein metric) and high correlation (Spearman’s rank sense) with other samples in the same class. Moreover, the newly defined spatial texture features consist of the Wasserstein distance between the optimally selected references and the remaining samples. We assessed the performance of the proposed classification pipeline in diagnosing the corona virus disease 2019 (COVID-19) from computed tomographic (CT) images.

## I. Introduction

Motivated by the goal of revealing the information hidden in standard-of-care images, the research field of radiomics has emerged [1]. Radiomics, which consists of the high-throughput extraction of quantitative features from medical images using automated quantification algorithms, is becoming a powerful tool in different applications, e.g., outcome prediction, tumor classification, treatment planning, and personalized therapy [2], [3], [4], [5] to cite a few examples. Among the computational radiomic quantifiers, texture is a key element that has been successful in different studies, e.g., [6], [7], [8], [9], although it does not have a universal adopted definition. Some authors proposed to define it as a measure of coarseness, contrast, directionality, line-likeness, regularity, and roughness [10].

Texture analysis may also be characterized as the quantification of spatial variations in gray levels within an image or region of interest (ROI). A variety of mathematical models have been proposed to evaluate the gray-level and the position of the pixels within an image [10]. One of the earliest methods using the spatial relationship to describe image texture features, which is known as the ***second-order histogram method*** and introduced in 1973 by Haralick *et al*., is the gray-level co-occurrence matrix (GLCM) [11]. The GLCM is defined as the joint probability of two pixels which have particular (cooccurring) gray-level values, with a distance *d* apart, and along a given direction *θ*. Another high-order statistic that characterizes local texture properties of an image in a similar manner as the GLCM approach, is the run-length matrix (RLM) [12]. A gray level run dictates the number of times two or more pixels having the same value in a preset direction, and the RLM is the matrix of run-length frequency occurring in an image in each considered direction. In radiomics research field, texture analysis consists usually of generating features based on summary statistics from high-dimensional feature matrices such as GLCM and RLM among others. This reduction step may lead to a loss of the spatial information that is inherent in these texture matrices, which has been discussed in previous studies [13], [14], [15]. The present paper is a continuation of a previous work highlighting the importance of spatial (multidimensional) texture features for robust medical image classification [15]. It is based on the assumption that there exist *representative* samples, which we refer to as *references* as well, i.e.,”good or bad” samples that represent each class/data cohort in a given particular classification/regression problem. This assumption results in new features which are represented as the distance between all remaining samples in the cohort and the selected reference samples. In particular, we propose to incorporate representative spatial texture features into a supervised image classification pipeline. The proposed spatial features are captured by the Wasserstein distance from optimal mass transport (OMT) theory [16], between the optimally selected reference GLCMs and the texture matrices of all other samples in the cohort. The selection of the reference samples is conducted in the training phase using a Bayesian optimization algorithm along with a support vector machine (SVM) classifier. A natural application of this advanced textural classification approach is in diseases with specific patterns. As an example, corona virus disease 2019 (COVID-19) related pneumonias, an infectious disease caused by the severe acute respiratory syndrome coronavirus 2 (SARS-CoV-2), have a prototypical radiological appearance that can be amenable to illustrate the virtues of the proposed methodology [17]. In this study, we test the performance of the supervised classification algorithm to effectively diagnose COVID-19 using computed tomographic (CT) images.

The increased interest in the use of OMT-based metrics, known as Wasserstein distance or Earth-Mover’s-Distance (EMD) in the engineering field, is mainly due to their natural ability to capture spatial information when comparing signals, images, or other types of data. This allows to provide various data distributions with different geometric interpretations, which we are seeking to capture from multidimensional texture matrices in the present work. In particular, the OMT problem seeks the most efficient way to transform one distribution of mass to another given a cost function [16]. Its origin goes back to 1781, when Gaspar Monge formulated the problem of finding the minimal transportation cost to redistribute earth for building fortifications [18]. Leonid Kantorovich in 1942, relaxed Monge’s formulation to find an optimal coupling of distributions using linear programming [19]. Since then, OMT has played a crucial role in many fields of science and engineering; see [20], [21] and references therein.

The remainder of the present paper is organized as follows. In Section II, the different components of the proposed classification pipeline are explained along with the data cohort and its pre-processing. Section III displays and discusses the results of the proposed classification algorithm, before concluding the paper in Section IV.

## II. Materials and Method

In this section, we will introduce the proposed classification algorithm and the data and application we investigated to test its performance. We tested the algorithm for diagnosing COVID-19 from CT images. At the time this paper is being written, the disease has infected more than 40 million individuals all over the world and caused more than 1 million deaths.

### A. Data and preprocessing

The COVID-19 CT images used in this study are publicly available^1^. The original dataset includes 349 CT scans that are positive for COVID-19 and 397 negative CT scans that are normal or contain other types of disease. The images were collected from 760 preprints about COVID-19. For further details about the collection process, we refer the interested reader to [22]. We resampled all images to a fixed grid of 512 × 512, and also equalized all the images to adapt their contrast. The lung masks have been derived based on a contrast stretching with an output range between *(−*1024, 226).

The lung fields were segmented using a modified UNet architecture as described here [23]. This network consists of encoding and decoding convolution blocks using a DenseNet design [24]. To avoid representation bottlenecks, the network adopts an Efficient Net (ENet) representation [25] that combines both max-pooling and strided convolutions on the same input. The architecture also uses a squeeze and excitation block in the encoders and decoder blocks to perform activation recalibration to enhance relevant features while suppressing irrelevant ones [26]. The network was trained with 2D slices from randomly selected set of CT scans spanning a range of pathologies including interstitial abnormalities that resembled some of the radiological patterns that can be found in patients with COVID-19 pneumonias. The pre-trained network is available in the Chest Imaging Platform open source toolkit [27]. This method was specifically designed to work without the need of volumetric data. After applying the lung segmentation pipeline in all images, non-valid segmentations were filtered out and removed which results in at the end 150 images for non-COVID patients and 174 images for COVID patients.

### B. Texture extraction

The GLCM is a two-dimensional matrix in which each element represents the frequency of occurrences of a pair of pixels in a spatial relation separated by a distance *d* and an angle *θ*. In our study, GLCM texture matrices were extracted automatically from the segmented lungs using the radiomics extension of the Computational Environment for Radiological Research (CERR) [28]. They were computed by combining contributions from all 2-D neighbors (i.e., *d* = 1 and *θ*_*i*_ = {0°, 45°, 90°, 135°}), using a gray quantization level value of 32. Additionally, 25 scalar statistical features, which are listed in Table I, were extracted from the GLCMs to be used in the proposed supervised classifier in addition to the newly defined spatial texture features.

**TABLE I:** GLCM-based scalar statistical features.

|  |  |  |
| --- | --- | --- |
| 1 : Energy | 10 : Inverse difference normalized | 18 : Cluster tendency |
| 2 : Joint entropy | 11 : Inverse variance | 19 : Cluster shade |
| 3 : Joint max | 12 : Difference entropy | 20 : Cluster prominence |
| 4 : Joint average | 13 : Difference variance | 21 : Haralick correlation |
| 5 : Joint variance | 14 : Sum average | 22 : Auto-correlation |
| 6 : Contrast | 15 : Sum entropy | 23 : First measure of information correlation |
| 7 : Inverse difference moment | 16 : Sum variance | 24 : Second measure of information correlation |
| 8 : Inverse difference moment normalized | 17 : Correlation | 25 : Dissimilarity |
| 9 : Inverse difference |  |  |

### C. Optimal mass transport and Wasserstein distance

We use the Wasserstein-1 distance from OMT theory [16] to capture the geometrical properties that are inherent in the GLCM texture matrices. The ***Wasserstein-1 distance*** between two *d*-dimensional probability distributions *ρ*_0_, *ρ*_1_ defined on Ω ⊂ ℝ^*d*^ is defined as follows:

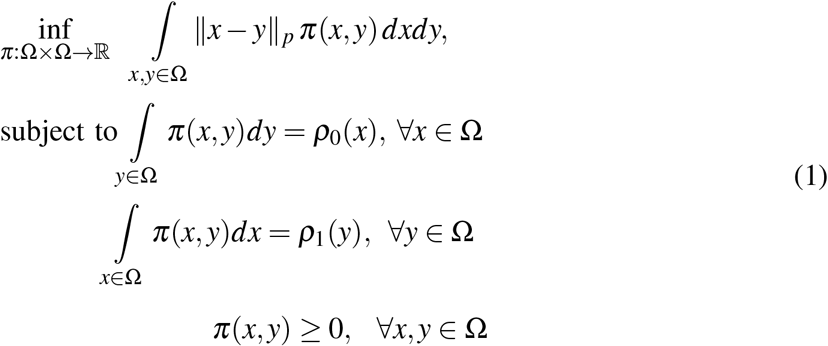

where ‖. ‖ _*p*_, 1 ≤ *p* ≤ ∞, is the ground metric of the Wasserstein distance. The variable *π* is the set of joint distributions *π*: Ω × Ω → ℝ whose marginal distributions are *ρ*_0_, *ρ*_1_.

An equivalent alternative formulation of the Wasserstein-1 distance, which is simpler and computationally more efficient, is defined by the following optimization problem:

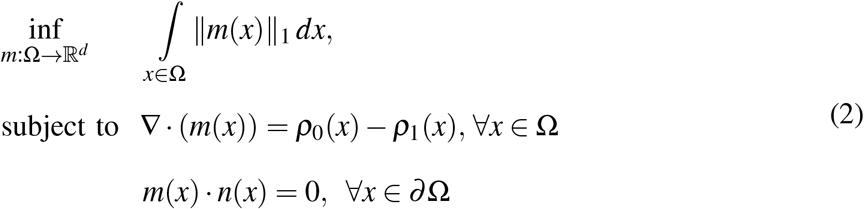

where *n*(*x*) denotes the normal to the boundary *∂* Ω, and *m* is a *d*-dimensional field satisfying the zero flux boundary condition [29]. A fast numerical scheme that relies on multilevel primal-dual optimization algorithms was proposed in [30] to solve (2). This latter numerical scheme is adopted in the present study.

### D. Selection of representative texture matrices and supervised image classification algorithm

In this paper, we refer to the good and bad samples in a given dataset as representative or reference samples. Moreover, the selection of such representative samples is presented for a binary classification task. However, it can be easily adopted in multi-label classification problems or even regression problems.

The main novelty of our approach consists of proposing a new set of features relying on Wasserstein distance, and the 2-D texture matrices which are considered as probability distributions. The main assumption that underlies the proposed set of features is that there may exist reference samples, either good or bad, in a studied dataset cohort (training set) and those references will be used to assess their similarity to the remaining training and testing texture samples through the Wasserstein metric.

Figure 1 depicts the devised workflow in our study. First, data were split into training (80%, 260 samples) and test (20%, 65 samples) sets. Then, reference samples are selected from each class of the training set as explained in the following. The procedure of learning and testing was replicated 100 times (Fig. 1).

**Fig. 1:**
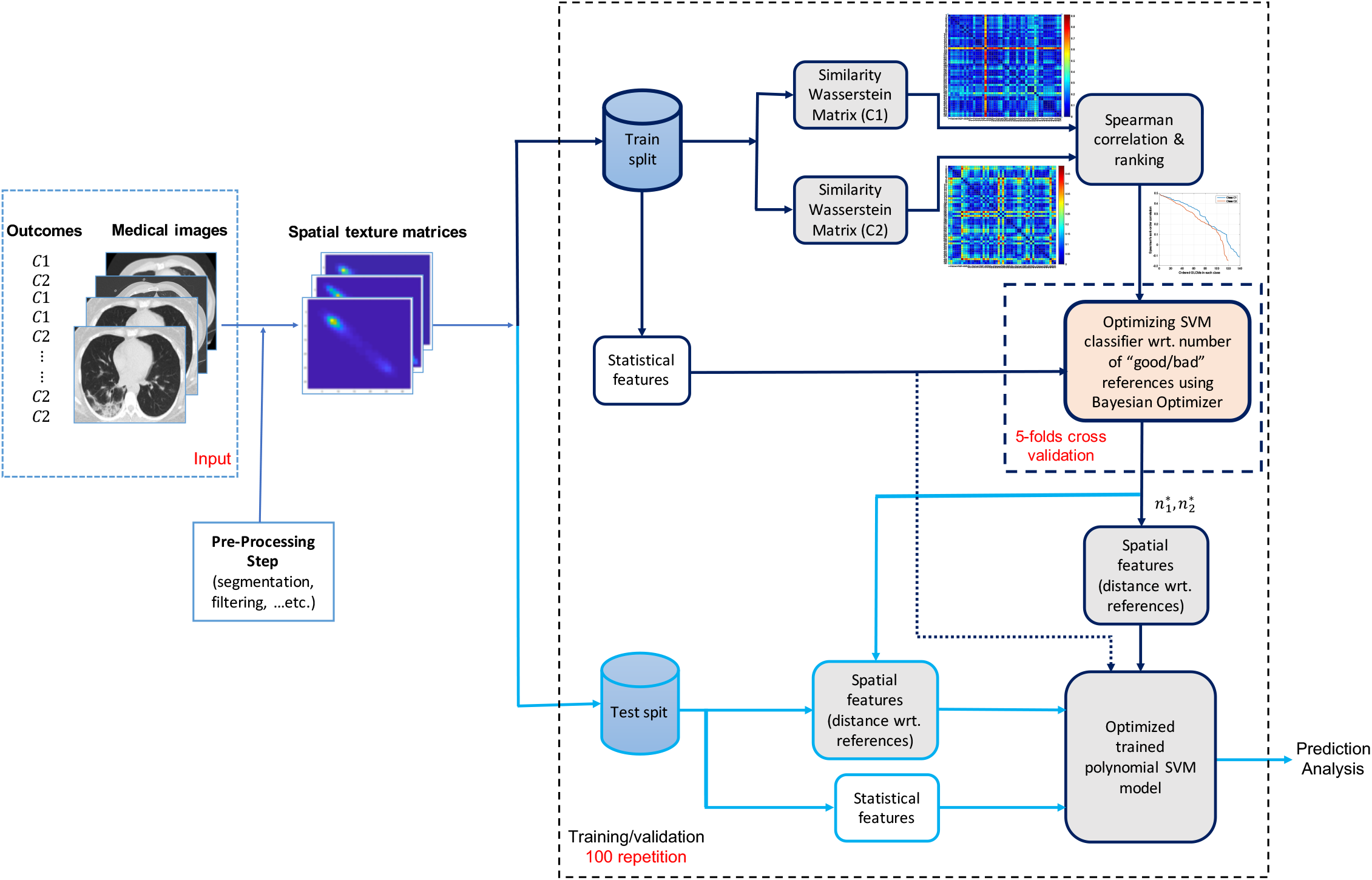
Flowchart of the proposed supervised classification algorithm using representative spatial texture features.

Let 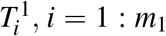, and 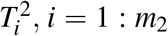, denote the GLCM texture matrices from the binary classes *C*_1_ and *C*_2_, respectively. We initially compute the Wasserstein-1 distance between all pairs 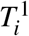 from *C*_1_ and all pairs 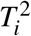 from *C*_2_ in the training set. The similarity measures are presented by the distance matrices 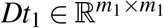 and 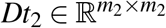. For each sample in a given class, we compute the correlation between its distance to all remaining samples in that class through the Spearman’s rank-order correlation given as follows:

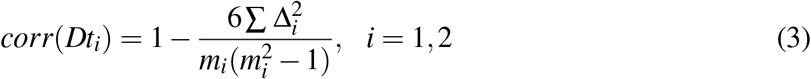

where Δ_*i*_, *i* = 1, 2 is the difference between the ranks of each pair of columns in *Dt*_1_ and *Dt*_2_, and *m*_*i*_, *i* = 1, 2 is the length of each column in *Dt*_1_ and *Dt*_2_.

Then, we order the average Spearman’s rank-order correlations of the GLCM samples 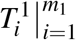 and 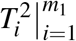. We refer to the good reference samples those having high average correlation ranks, and bad reference samples those with low average correlation ranks. Based on the computed averaged correlation ranks, we use Bayesian optimization algorithm to find the optimal number of good and bad samples (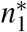 and 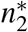) that maximizes the classification accuracy of the training set, or minimizes (1 *−accuracy*), as follows:

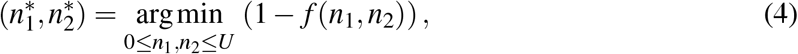

where *f* (*n*_1_, *n*_2_) is the training classification accuracy using *n*_1_ higher ranked samples and *n*_2_ lower ranked samples from the two classes *C*_1_ and *C*_2_. We note that the selected references are removed from the training set that is used to train the optimal SVM classifier. So implicitly, the proposed optimization approach optimizes the training efficiency, in addition to the training accuracy, which is captured through the number of training samples employed to build the prediction model after the reference samples have been removed. Moreover, the Wasserstein distance between the selected reference samples and each sample from the remaining training samples is added to the statistical features described previously. We should emphasize that the added Wasserstein metric based features between the reference and other training/testing samples captures the geometric similarity of their GLCM matrices to the good/bad representative GLCMs in each class. This is why we refer to these newly proposed features as *representative spatial texture features*.

A Bayesian optimizer has been chosen to find the optimal number of good/bad references in each class because of the advantages it offers. This class of optimization techniques is considered as a sequential design strategy that trades off exploration and exploitation mechanisms, unlike traditional active learning where the focus is often only in exploration, to attempt finding the global optimum of black-box functions, that do not usually assume any functional form, in a minimum number of steps [31]. This type of optimization approach has been adopted in different application fields [32]. It has been also very successful for hyper-parameters optimization when building predictive machine/deep learning-based models [33], [34].

## III. Results and Discussion

Our classification pipeline starts with assessing the similarity of all training GLCM pairs in each label class through the 1-Wasserstein distance. Figure 2 illustrates the similarity distance matrix of class 1 and class 2. As shown in Figure 2, some samples are closer in the sense of Wasserstein distance, compared to others, to the other samples within the same class. For instance, it is clear that sample 18 from class 1 is further away from other samples in that class compared to other samples. This observation has led us to the second step of our classification pipeline, which is to rank the similarity between samples within the same class through Spearman’s rank-order correlation. Then based on the ordered averaged correlation coefficient, we decide about the “good /bad” samples in a given class. Figure 3 shows the ranked correlation coefficients for all training samples in C1 and C2. We focus on the *n*_1_ samples from the tail of the ordered correlation vectors and *n*_2_ samples from its head. We optimized the accuracy of the trained SVM model, to select best values of *n*_1_ and *n*_2_, using Bayesian optimization and by performing 5-folds cross-validation. Given the optimal values of reference samples in each class we build an optimized classifier that accounts for the additional spatial texture features and that will serve for testing its accuracy using the “no seen” samples. The classification results are shown in Figure 4.

**Fig. 2:**
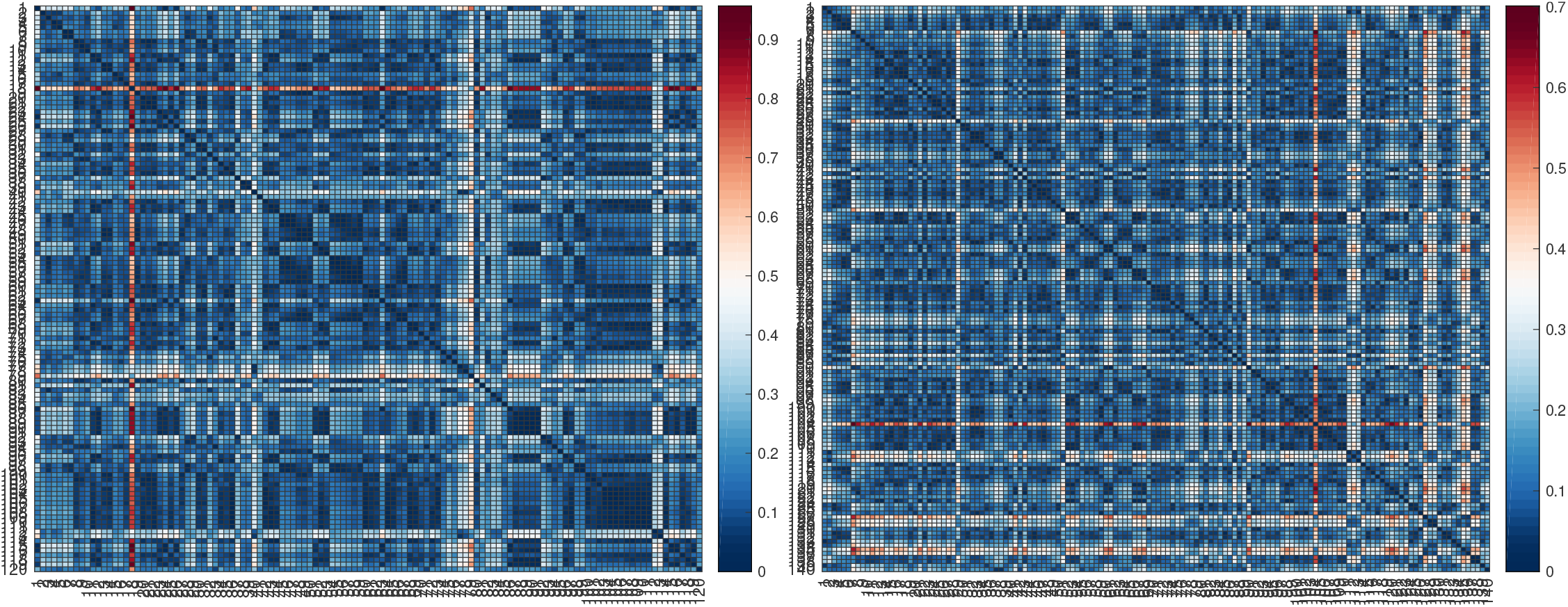
Distance similarity matrices between all training pairs of samples in Class 1 “*Dt*_1_” (left panel), and Class 2 “*Dt*_2_” (right panel).

**Fig. 3:**
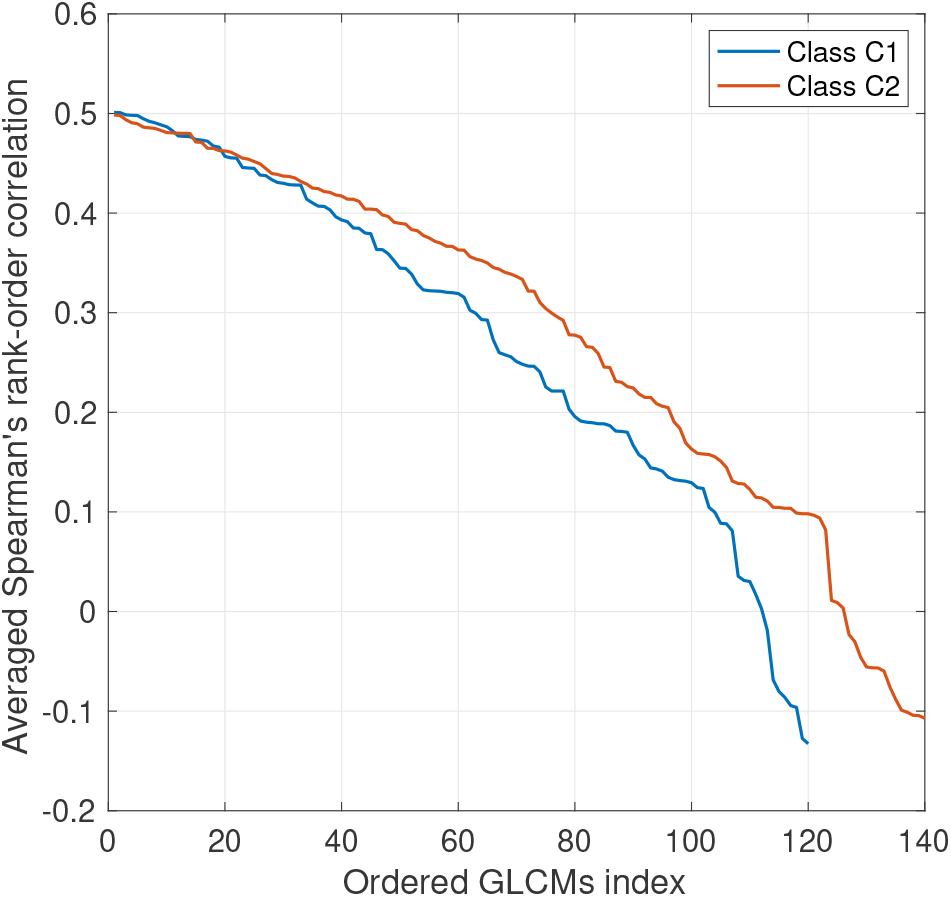
Averaged Spearman’s rank-order correlation for training Class 1 and Class 2.

**Fig. 4:**
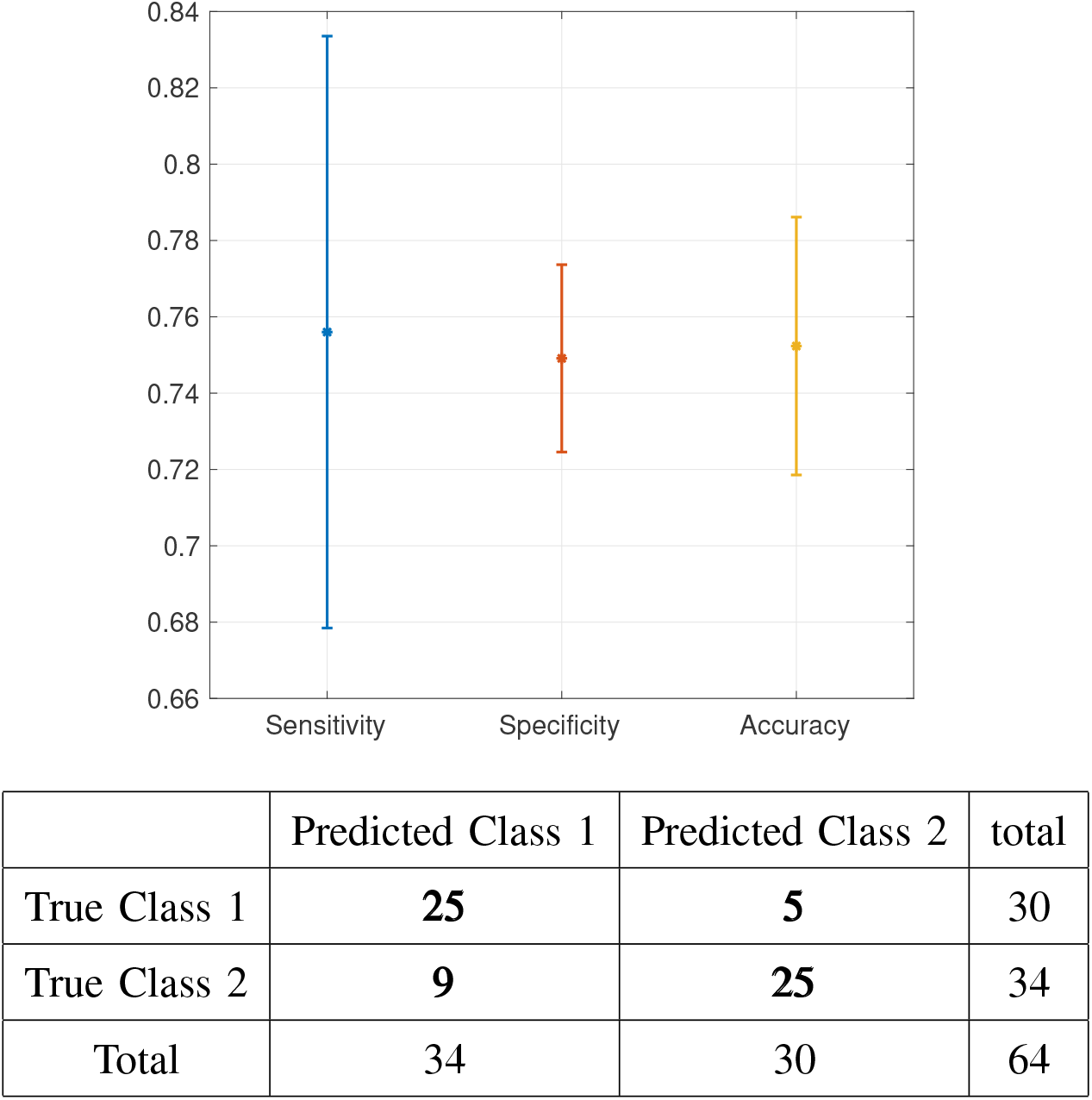
Classification performance for testing set. Upper panel: Classification statistics among 100 training/testing repetition, the mean value is highlighted with stars and standard deviation with bar plot. Lower panel: Confusion matrix for the maximum achieved classification accuracy.

The obtained results are very promising, especially given that the amount of data used to train the classifier is not large, and also given that the base classifier that is used (a third order polynomial SVM) does not inherently rely of many hyper-parameters, which may reduce the over-fitting problem that usually exists in prediction models that rely on deeper structures. Moreover, to better appreciate the added value of the newly introduced spatial GLCM features, we compared our proposed classifier with a third order polynomial SVM classifier that uses only the statistical features from GLCMs. It is worth-noting here that both the proposed SVM/OMT based classifier and the classical SVM classifier were trained/tested on the same training/test data cohorts, respectively. Prediction performance for both classifiers is provided in Table II. We observe from the table that the proposed classification pipeline outperforms the SVM classifier that is trained without the spatial texture features. This suggests the use of a subset of the training GLCMs as references of each class.

**TABLE II:** Classification results for test (“never seen”) data using the proposed classifier and classical SVM classifier. The **mean** and **std** represent the average and standard deviation of the classification statistics.

| Method | Proposed SVM/OMT based classifier<br>**mean (std)** | Polynomial SVM classifier<br>**mean (std)** |
| --- | --- | --- |
| Sensitivity | 0.7523 (0.0775) | 0.6603 (0.0614) |
| Specificity | 0.7491 (0.0245) | 0.7329 (0.0220) |
| Accuracy | 0.7560 (0.0338) | 0.6989 (0.0304) |

It is worth noting that in the present study, we only made use of the GLCM texture matrices to prove the concept of the additional value of the reference samples and the newly defined spatial texture features. Additional texture matrices, such as the RLM, can be included in the pipeline as well.

## IV. Conclusion

In this study, we proposed a classification pipeline for medical images that relies mainly on GLCM texture features together with Bayesian optimization and the Wasserstein-1 metric. The proposed method uses an optimal subset of samples to represent the training set and to define a new set of geometric texture features based on the given optimal subset. The results obtained indicated the importance of the reference sample selection step and consequently the newly defined spatial features in the classification work flow. The present work provides a way forward to more extensive studies with more data in order to build efficient prediction methods for analyzing medical images for various clinical scenarios.

## Data Availability

The data is publicly available

## Acknowledgments

This study was supported in part by AFOSR grants (FA9550-17-1-0435, FA9550-20-1-0029), NIH grant (R01-AG048769), MSK Cancer Center Support Grant/Core Grant (P30 CA008748), and a grant from Breast Cancer Research Foundation (grant BCRF-17-193).

The authors would like to thank Prof. Marc Niethammer for the discussions about lung segmentation, Prof. Joseph Deasy for the fruitful discussions about the proposed classification algorithm, and Dr. Saad Nadeem for discussions about texture analysis and the data used in this study.

## Footnotes

1 https://github.com/UCSD-AI4H/COVID-CT

## References

[1] P. Lambin, E. Rios-Velazquez, R. Leijenaar, and et al., “Radiomics: extracting more information from medical images using advanced feature analysis,” European Journal of Cancer, vol. 48, pp. 441–446, 2012.

[2] O. Morin, M. Vallières, A. Jochems, and et al., “A deep look into the future of quantitative imaging in oncology: A statement of working principles and proposal for change,” International Journal of Radiation Oncology, vol. 102, pp. 1074–1082, 2018.

[3] J. C. Peeken, M. Bernhofer, B. Wiestler, and et al., “Radiomics in radiooncology-challenging the medical physicist,” European Journal of Medical Physics, vol. 48, pp. 27–36, 2018.

[4] P. Lambin, R. T. Leijenaar, T. M. Deist, and et al., “Radiomics: the bridge between medical imaging and personalized medicine,” Nature Reviews Clinical Oncology, vol. 14, pp. 749–762, 2017.

[5] R. Shiradkar, T. K. Podder, A. Algohary, and et al., “Radiomics based targeted radiotherapy planning (Rad-TRaP): a computational framework for prostate cancer treatment planning with MRI,” Radiation Oncology, vol. 11, pp. 1–14, 2016.

[6] J. Meng, S. Liu, L. Zhu, and et al., “Texture analysis as imaging biomarker for recurrence in advanced cervical cancer treated with CCRT,” Scientific reports, vol. 8, pp. 1–9, 2018.

[7] Z. Yan, J. Zhang, H. Long, and et al., “Correlation of CT texture changes with treatment response during radiation therapy for esophageal cancer: An exploratory study,” PLoS ONE, vol. 14, pp. 1–12, 2019.

[8] E. Scalco and G. Rizzo, “Texture analysis of medical images for radiotherapy applications,” British institute of radiology, vol. 90, pp. 1–15, 2016.

[9] A. Ahmed, P. Gibbs, M. Pickles, and L. Turnbull, “Texture analysis in assessment and prediction of chemotherapy response in breast cancer,” J Magn Reson Imaging, vol. 38, pp. 89–101, 2013.

[10] C. Hung, E. Song, and Y. Lan, Image texture analysis: Foundations, models and algorithms. Springer International Publishing, 2019.

[11] R. M. Haralick, K. Shanmugam, and I. Dinstein, “Textural features for image classification,” IEEE Transcations on Systems, Man and Cybernetics, vol. 3, pp. 610–621, 1973.

[12] M. M. Galloway, “Texture analysis: A review of neurologic MR imaging applications,” Comput. Graph. Image Process., vol. 4, pp. 172–179, 1975.

[13] A. L. Vickers and J. W. Modestino, “A maximum likelihood approach to texture classification,” IEEE Transactions on Pattern Analysis and Machine Intelligence, vol. PAMI-4, pp. 61–68, 1982.

[14] X. Li, M. Guindani, C. S. Ng, and B. P. Hobbs, “Spatial bayesian modeling of glcm with application to malignant lesion characterization,” Journal of applied statistics, vol. 46, pp. 230–246, 2019.

[15] Z. Belkhatir, A. Iyer, J. C. Mathews, M. Pouryahya, S. Nadeem, J. O. Deasy, A. P. Apte, and A. R. Tannenbaum, “Optimal mass transport for robust texture analysis,” Available at: https://www.biorxiv.org/content/10.1101/855221v1.full.pdf, 2020.

[16] C. Villani, Optimal transport: old and new. Springer-Verlag Berlin Heidelberg, 2009.

[17] A. Kaufman, S. Naidu, S. Ramachandran, D. Kaufman, Z. Fayad, and V. Mani, “Review of radiographic findings in COVID-19,” World Journal of Radiology, vol. 12, pp. 142–155, 2020.

[18] G. Monge, Mémoire sur la théorie des déblais et des remblais. De l’Imprimerie Royale, 1781.

[19] L. V. Kantorovich, “On the translocation of masses,” Journal of mathematical sciences, vol. 133, 2006.

[20] L. C. Evans, “Partial differential equations and monge-kantorovich mass transfer,” in Current Developments in Mathematics, International Press, 1999.

[21] S. Kolouri, S. Park, M. Thorpe, and et al., “Optimal mass transport: signal processing and machine-learning applications (survey paper),” IEEE Signal Processing Magazine, vol. 34, pp. 43–59, 2017.

[22] X. He, X. Yang, S. Zhang, J. Zhao, Y. Zhang, E. Xing, and P. Xie, “Sample-efficient deep learning for COVID-19 diagnosis based on CT scans,” Available at: https://www.medrxiv.org/content 10.1101/2020.04.13.20063941v1.full.pdf, 2020.

[23] R. Moreta-Martz, G. V. Shez-Ferrero, L. Andresen, J. Q. Holsting, and R. S. J. Estépar, “Multi-cavity heart segmentation in non-contrast non-ECG gated CT scans with F-CNN,” in Thoracic Image Analysis. TIA 2020. Lecture Notes in Computer Science, J. Petersen and et al., Eds., vol. 12502. Springer, Cham, 2020, pp. 2951–2959.

[24] G. Huang, Z. Liu, L. van der Maaten, and K. Q. Weinberger, “Densely connected convolutional networks,” IEEE Conference on Computer Vision and Pattern Recognition (CVPR), p. 2261–2269, 2017.

[25] A. Paszke, A. Chaurasia, S. Kim, and E. Culurciello, “ENet: A deep neural network architecture for real-time semantic segmentation,” Available at: https://arxiv.org/pdf/1606.02147.pdf, 2016.

[26] A. G. Roy, N. Navab, and C. Wachinger, “Concurrent spatial and channel squeeze & excitation in fully convolutional networks,” Available at: https://arxiv.org/pdf/1803.02579.pdf, 2018.

[27] R. S. J. Estépar, J. Ross, R. Harmouche, J. Onieva, A. Diaz, and G. Washko, “Chest imaging platform: An open-source library and workstation for quantitative chest imaging,” American Thoracic Society International Conference Abstracts American Thoracic Society, p. A4975?A4975, 2015.

[28] A. P. Apte and et al., “Extension of CERR for computational radiomics: A comprehensive matlab platform for reproducible radiomics research,” Medical physics, vol. 45, pp. 3713–3720, 2018.

[29] M. Beckmann, “A continuous model of transportation,” Econometrica: Journal of the Econometric Society, vol. 129, pp. 643–660, 1952.

[30] J. Liu, W. Yin, W. Li, and Y. T. Chow, “Multilevel optimal transport: A fast approximation of Wasserstein-1 distances,” Available: https://arxiv.org/pdf/1810.00118.pdf, 2018.

[31] P. I. Frazier, “A tutorial on bayesian optimization,” Available at: https://arxiv.org/pdf/1807.02811.pdf, 2018.

[32] B. Shahriari, K. Swersky, Z. Wang, R. P. Adams, and N. de Freitas, “Taking the human out of the loop: A review of bayesian optimization,” Proceedings of the IEEE, vol. 104, pp. 148–175, 2016.

[33] J. Snoek, H. Larochelle, and R. P. Adams, “Practical bayesian optimization of machine learning algorithms,” in Advances in Neural Information Processing Systems, F. Pereira, C. J. C. Burges, L. Bottou, and K. Q. Weinberger, Eds., vol. 25. Curran Associates, Inc., 2012, pp. 2951–2959.

[34] F. Archetti and A. Candelieri, Bayesian optimization and data science. Springer Briefs in Optimization, 2019.

